# General healthcare preferences for people aged 50 and over in England: English Longitudinal Study of Ageing

**DOI:** 10.64898/2026.01.18.26344363

**Authors:** Nicholas Steel, Paola Zaninotto, Oby O. Enwo, James Banks

## Abstract

**Background:** Treatment outcomes and satisfaction improve when healthcare decisions align with patient preferences. However, little is known about general health preferences in older adults, and eliciting patient preferences is time consuming.

**Aim:** To describe the distribution of preferences for healthcare in general, and associations between preferences and patient characteristics in adults aged 50 years and over in England.

**Design and setting:** Data were collected in wave 8 (2016/2017) of the English Longitudinal Study of Ageing (ELSA), a biennial longitudinal survey of people aged 50 and over living in private households in England that began in 2002/2003 and is designed to be nationally representative.

**Method:** Data were collected from ELSA participants using face-to-face interviews and a short self completion questionnaire. Amongst other items, the questionnaire contained six new measures of general healthcare preferences: risk aversion, future orientation, quality or length of life, body function compared to looks, openness to experimental treatments, and preference to leave treatment decisions to a doctor.

**Results:** Healthcare preferences varied substantially, with some systematic patterns by age and sex: women were less likely than men to want to delegate decisions to doctors, and more likely to prioritise quality of life over length of life; participants aged over 75 years were more likely than younger participants to want to delegate decisions to doctors, and more likely to want to avoid risks.

**Conclusion:** The systematic sex and age differences offer a useful starting point for eliciting healthcare preferences in a busy clinical environment, but cannot replace asking about individual preferences.

## Introduction

Healthcare decisions have traditionally been primarily guided by clinical expertise and standardized guidelines with limited patient input, and this approach is still often appropriate, particularly for acute conditions.[1] However, shared decision making (SDM), where clinicians and patients collaboratively make healthcare decisions using the best evidence alongside patient preferences, has been widely advocated for at least 20 years for two main reasons: the ethical imperative to involve patients with their care, and the evidence that delivering care that resonates with patient preferences can both improve outcomes and reduce costs, for example by reducing rates of surgery.[2–5] Patient preferences for health care encompass personal values, priorities, and trade-offs regarding the risks, benefits, and side effects of treatment options.

Progress with integrating patient preferences into routine healthcare decision-making has been slow, despite almost universal agreement about its importance and the development of many high quality decision aids for specific conditions, designed to help patients understand complex medical information and consider the trade-offs between different options.[6–8] However, few decision aids have been developed for older adults, who may have frequent contact with healthcare services and often face complex healthcare decisions.[8] Individual preferences are inherently diverse and little is known about the extent to which population groups may share general health preferences for healthcare, as opposed to preferences about treatment for a specific condition.

More evidence is needed on general healthcare preferences, particularly among older adults. This study achieved its aim to explore the distribution of general healthcare preferences and associations with patient characteristics among adults aged 50 years and over in England, using a specially designed short questionnaire delivered to participants in Wave 8 of the English Longitudinal Study of Ageing (ELSA).

## Materials and Methods

The English Longitudinal Study of Ageing (ELSA) is an open-access biennial longitudinal survey of those aged 50 and over living in private households in England that began in 2002/2003.[9] The data set is available from the UK Data Service.[10] The sample was drawn from participants in the Health Survey for England (HSE), an annual cross-sectional survey that was designed to monitor the health of the general population. Data were collected through face-to-face interviews and self-completion questionnaires. Comparisons of the socio-demographic characteristics of participants against results from the 2011 national census indicate that the sample was broadly representative of the English population.

### Development of general healthcare preference items

A literature search identified questionnaires that covered autonomy, shared decision making and beliefs about medication, but no suitable existing measure of general healthcare preferences.[11–15] The literature review found five relevant questionnaires which covered autonomy, shared decision making and beliefs about medications, and in the absence of an existing measure of general health preferences, we modified concepts from these scales and from Mulley[16], informed by established ELSA questions on attitudes to financial risk, to develop six questions about general healthcare preferences for use in ELSA. The questions covered risk aversion, future orientation, quality or length of life, body function compared to looks, openness to experimental treatments, and preference to leave treatment decisions to a doctor (Table 2).

These new questions were asked for the first time to all participants in wave 8 of ELSA (2016/2017). The questionnaire is freely available.[17] The questions were asked in the paper-based self-completion questionnaire (question numbers 56–61), which was either returned by the interviewer or posted back by the respondent in a Freepost envelope provided by the interviewer.[18] The questions followed on from the similar established questions about attitudes to financial risk. Each item was assessed on a scale from 0 to 10, with 10 indicating a preference for low risk. The order of responses for the first question (avoid risks) was reversed for all analyses in this paper, so as to be consistent with 10 indicating a preference for lower risk. This question was originally asked with 0 being ‘Avoid taking risks’ and 10 being ‘Fully prepared to take risks’. The remaining 5 questions were analysed with the response order unchanged from that in the questionnaire.

### Data

Demographic characteristics were recorded from the main questionnaire, with the exception of alcohol which was asked about in the self-completion questionnaire. Age was categorised into three groups (50-64, 65-74 and over 75 years). Sex was categorised as male or female. Education was categorized into high (College/University and above), intermediate (Advanced-level) and low (Ordinary-level or lower). Region of residence was categorised into 9 English regions. Coronary heart disease (CHD), diabetes, respiratory disease, stroke and cancer were assessed using self-reported doctor diagnosis. Smoking status was classified as never smoked, ex-smoker or current smoker. Respondents asked whether they ‘take part in sports or activities that are vigorous / moderately energetic / mildly energetic’ in 3 separate questions. A binary variable was created for ‘active or ‘inactive’ by classifying participants who reported doing vigorous or moderate physical activity at least once per week as ‘active’, and those who reported doing both vigorous and moderate activity less often than once per week as ‘inactive’. The frequency of drinking alcoholic drinks over the last 12 months was classified into ‘daily’ (almost every day) or ‘less than daily’ (five or six days a week or less, including no alcoholic drinks in the last 12 months).

ELSA was conducted in accordance with the Declaration of Helsinki. ELSA Wave 8 received ethical approval from the South Central – Berkshire Research Ethics Committee on 23rd September 2015 (15/SC/0526). All ELSA participants provided written consent to take part in the study. Data are made available through the UK Data Service.

### Statistical analysis

Descriptive statistics (means, percentages and 95% CIs) and statistical analyses were estimated using the wave 8 self-completion weight (w8scwt), accounting for clustering (idahhw8) and stratification by government office region (gor). The ELSA wave 8 self-completion weight was a product of a non-response weight designed to minimise any bias arising from differential non-response to the self-completion questionnaire, and the wave 8 cross-sectional weight (scaled so that the average weight was equal to 1). Linear regression models were used to estimate the fraction of variance explained by different clusters of covariates, and the association between each health preference item and the full set of included covariates. Principal component analysis was used to explore whether the health preference items could be represented by fewer underlying core components. ‘Don’t know’ values were recoded as missing. All analyses were run in Stata SE15 and R version 4.5.2.

### Sensitivity analyses

The impact of missing data on the results was tested with a sensitivity analysis. We tested whether the prevalence of variance explained by each predictor was similar when we restricted the analyses to the 4,727 respondents with valid answers on all items and no missing data on predictors. A non-negligible proportion of the sample selected the middle score of 5 on almost all items, perhaps capturing people who did not know, rather than people who wanted to report a central value. Thus we also re-ran all analyses excluding responses reporting 5, to check whether results were driven by this score. Lastly, we calculated odds ratios from logistic regressions for the likelihood of responding 5 vs not 5, and for the likelihood of strong positives and strong negatives for each healthcare preference item.

## Results

7,133 core ELSA members responded to wave 8 main interview (response rate 82.4%) and of these, 6,257 responded to the self-completion questionnaire, 6,133 core members completed at least one healthcare preference item, and the final weighted sample was 6,098. The mean age was 66 years, with slightly more women (52%) than men. 33% had low education, 11% smoked, 23% were physically inactive, and 12% drank alcohol daily. Most (72%) reported having none of our five included chronic conditions (CHD, diabetes, respiratory disease, stroke, and cancer); conditions commonly reported included respiratory disease (13%) and diabetes (11%) (Table 1).

**Table 1.** Demographic and health characteristics of participants, English Longitudinal Study of Ageing self completion questionnaire, wave 8.

| **Characteristic** | **%** | **Low**<br>**95%CI** | **High 95%CI** |
| --- | --- | --- | --- |
| Male | 47.6 | 46.1 | 49.1 |
| Female | 52.4 | 50.9 | 53.9 |
| Mean Age (years) | 66.4 | 66.0 | 66.8 |
| 50-64 | 47.6 | 45.7 | 49.5 |
| 65-74 | 29.9 | 28.5 | 31.4 |
| 75 and older | 22.5 | 21.2 | 23.8 |
| **Education** |  |  |  |
| High | 19.6 | 18.2 | 21.1 |
| Intermediate | 47.1 | 45.3 | 48.9 |
| Low | 33.3 | 31.7 | 34.8 |
| **Region** |  |  |  |
| South East | 17.1 | 16.1 | 18.1 |
| London | 11.6 | 10.7 | 12.6 |
| North East | 5.2 | 4.8 | 5.6 |
| North West | 13.2 | 12.4 | 14.1 |
| Yorkshire and The Humber | 10.1 | 9.4 | 10.9 |
| East Midlands | 9.0 | 8.5 | 9.7 |
| West Midlands | 10.6 | 10.0 | 11.3 |
| East of England | 11.7 | 11.0 | 12.5 |
| South West | 11.4 | 10.7 | 12.2 |
| **Smoking Status** |  |  |  |
| Never smoked | 39.3 | 37.6 | 41.1 |
| Ex-smoker | 49.8 | 48.0 | 51.6 |
| Current smoker | 10.9 | 9.7 | 12.1 |
| **Frequency of alcohol consumption** |  |  |  |
| Less than daily (6 or less days a week) | 87.7 | 86.6 | 88.8 |
| Daily (7 days a week) | 12.3 | 11.2 | 13.4 |
| **Physical activity** |  |  |  |
| Active | 77.3 | 75.9 | 78.7 |
| Inactive | 22.7 | 21.3 | 24.1 |
| **Chronic conditions** |  |  |  |
| Coronary heart disease | 5.0 | 4.4 | 5.5 |
| Diabetes | 10.5 | 9.6 | 11.5 |
| Respiratory | 13.0 | 11.9 | 14.2 |
| Stroke | 3.3 | 2.8 | 3.8 |
| Cancer | 3.2 | 2.7 | 3.8 |
| **Number of chronic conditions** |  |  |  |
| None | 71.7 | 70.2 | 73.2 |
| 1 | 22.7 | 21.3 | 24.1 |
| 2+ | 5.6 | 4.9 | 6.3 |
| **Total N (weighted)** | **6,098** |  |  |

There was an approximately even distribution of responses across the full range for 4 of the 6 healthcare preference questions: avoid risks, live for the future, avoid experimental treatments, and leave treatment decisions to doctor (Table 2, Fig 1). The responses to the question on length of life had higher scores for quality of life as opposed to length of life, and the question on body function had consistently higher scores for body function as opposed to body looks. The question on body function was the only item where response 5, the midpoint of the scale, was not the highest scoring response. Response category 11 (‘Don’t know’) was below 5% for all items except for the question on leaving treatment decisions to doctor, where it was over 10%.

**Figure 1.**
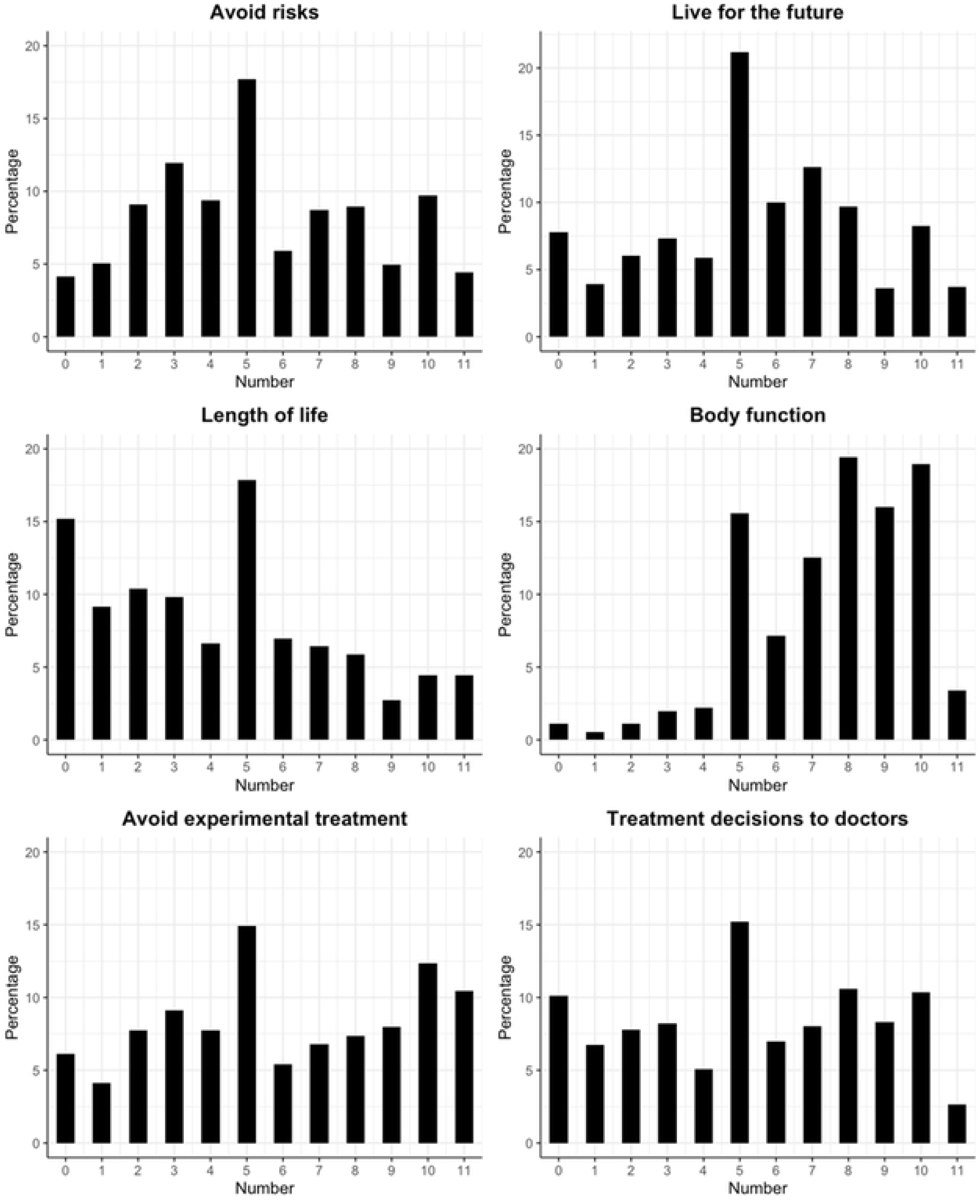
Distribution of responses to health preference questions. Each item was assessed on a scale from O to 10, with 10 indicating a preference for low risk (see Table 2 for question wording and response categories). 11 represents ‘Don’t know’ responses.

**Table 2.** Responses to health preference questions.

| Question | N Weighted | Mean | Std. Dev. | Value | % |
| --- | --- | --- | --- | --- | --- |
| **1 Avoid risks**<br>Thinking specifically about the risks and benefits of treatments to improve your health, are you a person who is fully prepared to take risk or do you try to avoid taking risks?<br>[0 'Fully prepared to take risks'...10 'Avoid taking risks'] | 5,793 | 5.2 | 2.8 | 0 -1<br>2-3<br>4-5<br>6-7<br>8-9<br>10 | 9.6<br>22.0<br>28.3<br>15.3<br>14.6<br>10.2 |
| **2 Live for the future**<br>Thinking about your health in general. Do you generally prefer to live for the moment, or to live for the future?<br>[0 'Live for the moment'...10 'Live for the future'] | 5,838 | 5.3 | 2.8 | 0 -1<br>2-3<br>4-5<br>6-7<br>8-9<br>10 | 12.2<br>13.9<br>28.1<br>23.5<br>13.8<br>8.5 |
| **3 Length of life important**<br>Is quality of life generally more important to you, or length of life?<br>[0 'Quality of life'...10 'Length of life'] | 5,793 | 4.0 | 2.9 | 0 -1<br>2-3<br>4-5<br>6-7<br>8-9<br>10 | 25.5<br>21.2<br>25.6<br>14.1<br>9.0<br>4.7 |
| **4 Body function important**<br>Is how your body looks usually more important to you, or how your body functions?<br>[0 'How my body looks'...10 'How my body functions'] | 5,862 | 7.4 | 2.2 | 0 -1<br>2-3<br>4-5<br>6-7<br>8-9<br>10 | 1.7<br>3.2<br>18.4<br>20.4<br>36.7<br>19.6 |
| **5 Avoid experimental treatments**<br>Are you a person who would be willing to use experimental treatments, or someone who would never use experimental treatments?<br>[0 'Willing to use experimental treatments'...10 'Would never use experimental treatments'] | 5,429 | 5.4 | 3.1 | 0 -1<br>2-3<br>4-5<br>6-7<br>8-9<br>10 | 11.4<br>18.8<br>25.3<br>13.6<br>17.1<br>13.8 |
| **6 Leave treatment decisions to doctor**<br>Are you a person who prefers to make the final decision about your healthcare, or someone who prefers to leave all treatment decisions to their doctor?<br>[0 'Prefer to make the final treatment decision' 10 'Prefer to leave all treatment decisions to my doctor'] | 5,913 | 5.2 | 3.2 | 0 -1<br>2-3<br>4-5<br>6-7<br>8-9<br>10 | 17.3<br>16.4<br>20.8<br>15.4<br>19.4<br>10.6 |

Pearson’s correlation coefficients quantified the relationships between healthcare preferences (Table 3). ‘Live for the future’ and ‘length of life’ correlated negatively with ‘avoid risks’, indicating that risk-averse individuals valued quality of life at the present time more highly than in the future. Avoiding risks correlated positively with avoiding experimental treatments and deferring decisions to doctors.

**Table 3.** Correlations between healthcare preference questions (Pearson correlation coefficients)

| Question | 1 | 2 | 3 | 4 | 5 | 6 |
| --- | --- | --- | --- | --- | --- | --- |
| 1 Avoid risks | 1 |  |  |  |  |  |
| 2 Live for the future | -0.11 | 1 |  |  |  |  |
| 3 Length of life important | -0.05 | 0.35 | 1 |  |  |  |
| 4 Body function important | -0.06 | 0.11 | -0.02 | 1 |  |  |
| 5 Avoid experimental treatments | 0.25 | 0.06 | 0.12 | 0.02 | 1 |  |
| 6 Leave treatment decisions to doctor | 0.02 | 0.11 | 0.23 | 0.03 | 0.12 | 1 |

Principal component analysis indicated that the first component accounted for just over one quarter (0.257) of the total variance, and a four-component solution accounted for just over three quarters (0.783) of the total variance (Table 4). The six healthcare preference items clustered into four distinct components, showing the multidimensional nature of healthcare preferences.

**Table 4.**
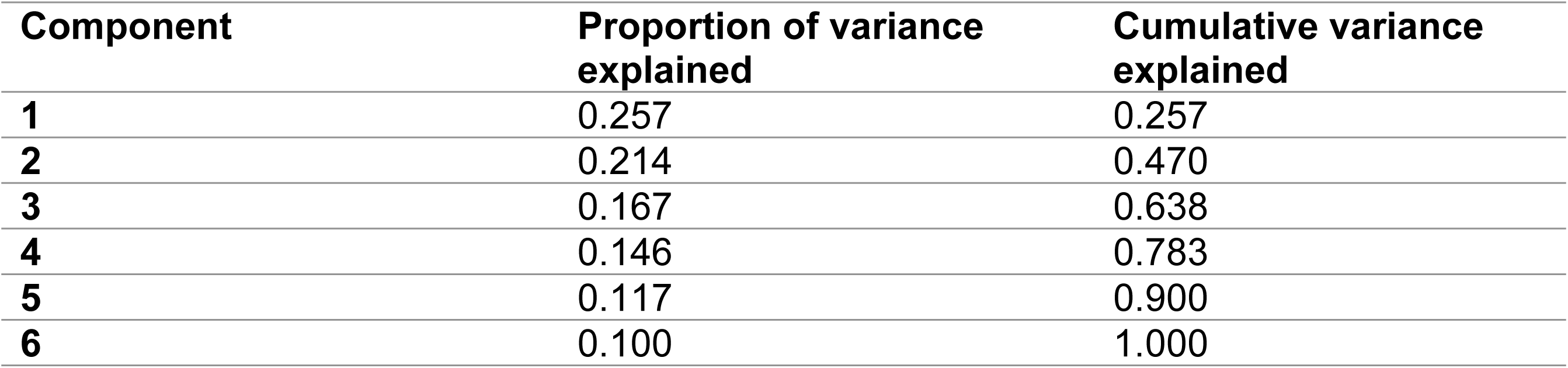
Principal component analysis of health preference questions.

| Component | Proportion of variance explained | Cumulative variance explained |
| --- | --- | --- |
| 1 | 0.257 | 0.257 |
| 2 | 0.214 | 0.470 |
| 3 | 0.167 | 0.638 |
| 4 | 0.146 | 0.783 |
| 5 | 0.117 | 0.900 |
| 6 | 0.100 | 1.000 |

Older individuals, particularly those aged 75 years or older, were more likely than those aged between 50 and 64 to want to avoid risks (linear regression coefficient 0.470, p<0.001), prioritize body function over looks (0.558, p<0.001), avoid experimental treatments (0.759, p<0.001), and leave treatment decisions to doctors (0.880, p<0.001) (Table 5). Women were more likely than men to want to avoid risks (0.443, p<0.001), prioritize quality of life over length of life (-0.566, p<0.001), prioritize body looks over body function (-0.298, p<0.001), avoid experimental treatments (0.652, p<0.001), and were less likely to want to defer treatment decisions to doctors (-0.732, p<0.001). Lower educational attainment was associated with wanting to avoid risks (0.391, p<0.01), prioritize length over quality of life (0.680, p<0.001), avoid experimental treatments (0.506, p<0.01), and leave treatment decisions to doctors (1.481, p<0.001).

**Table 5.** Associations between participant characteristics and health preferences (linear regression coefficients)

| Covariates | Avoid Risks | Live for the future | Length of life | Body Function | Avoid experimental treatment | Treatment decisions to doctors |
| --- | --- | --- | --- | --- | --- | --- |
| (Intercept) | 5.157 (***) | 5.793 (***) | 4.383 (***) | 7.288 (***) | 5.523 (***) | 4.437 (***) |
| **Age 50-64 (baseline)** |  |  |  |  |  |  |
| **Age 65-74** | 0.036 NS | -0.093 NS | -0.074 NS | 0.303 *** | -0.024 NS | 0.162 NS |
| **Age 75+** | 0.470 *** | 0.045 NS | 0.111 NS | 0.558 *** | 0.759 *** | 0.880 *** |
| **Female (vs male)** | 0.443 *** | -0.165 | -0.566 *** | -0.298 *** | 0.652 *** | -0.732 *** |
| **High Education (baseline)** |  |  |  |  |  |  |
| **Intermediate Education** | 0.009 NS | -0.229 NS | 0.169 NS | -0.076 NS | 0.184 NS | 0.716 *** |
| **Low Education** | 0.391 ** | -0.129 NS | 0.680 *** | 0.009 NS | 0.506 ** | 1.481 *** |
| **Never smoked (baseline)** |  |  |  |  |  |  |
| **Ex-Smoker** | -0.152 NS | -0.001 NS | -0.058 NS | -0.250 ** | -0.309 * | -0.131 NS |
| **Current Smoker** | 0.062 NS | -0.943 *** | -0.164 NS | -0.204 NS | -0.282 NS | -0.019 NS |
| **Alcohol daily (vs less than daily)** | 0.134 NS | -0.171 NS | -0.164 NS | 0.099 NS | -0.359 * | -0.435 ** |
| **Physical inactivity** | 0.297 * | -0.167 NS | -0.301 * | 0.231 * | 0.202 NS | 0.261 NS |
| **Coronary heart disease** | -0.754*** | 0.030 NS | -0.149 NS | -0.131 NS | -0.384 NS | -0.232 NS |
| **Diabetes** | 0.019 NS | 0.165 NS | 0.228 NS | 0.192 NS | -0.308 NS | 0.209 NS |
| **Respiratory Disease** | -0.143 NS | -0.146 NS | 0.335 NS | -0.003 NS | -0.107 NS | 0.031 NS |
| **Stroke** | -0.018 NS | -0.056 NS | -0.274 NS | -0.087 NS | -0.145 NS | 0.549 * |
| **Cancer** | -1.040 *** | -0.274 NS | 0.079 NS | 0.160 NS | -0.393 NS | -0.974 ** |
| **Total N** | 5586 | 5632 | 5593 | 5653 | 5256 | 5706 |

\* p<0.05, \*\* p<0.01, \*\*\* p<0.001, NS = no statistically significant difference. Total N reflects participants with valid responses for the health preference item, all covariates, and non-zero survey weights.

Ex-smokers were less likely than never smokers to want to prioritise body looks over body function (-0.250, p<0.01), and current smokers wanted to live for the moment rather than for the future (-0.943, p<0.001). Daily alcohol drinkers were less likely to leave treatment decisions to doctors (-0.435, p<0.01). Individuals with cancer were less likely to want to avoid risks (-1.040, p<0.001). The full results for each individual healthcare preference showed little variation by region (Supplementary Tables S1-S6).

### Sensitivity Analysis

The percentage of variance explained by each predictor was similar when we restricted the analyses to participants with valid answers on all items and no missing data on predictors (n=4,727) (Supplementary Table S7). Associations between participant characteristics and healthcare preferences were also similar (Supplementary Table S8).

We re-ran the linear regression analyses after excluding responses reporting 5, to check whether results were driven by this score. When responses with a score of 5 were removed, the direction and overall pattern of associations remained similar with the main analysis (Supplementary Table S9). This indicates that the main analysis results are not driven by midpoint responding.

The odds ratios from logistic regressions for the likelihood of responding 5 vs not 5, and for the likelihood of strong positives and strong negatives for each healthcare preference item showed selective associations across certain covariates. Older adults were less likely to select 5 (vs not 5) for ‘Body function’ (odds ration 0.581, p<0.001), however overall patterns did not indicate systematic covariate-specific inflation of mid-point responses (Supplementary Table S10).

Women were more likely to provide strong positive responses for ‘avoid risks’ and ‘avoid experimental treatment’ (Supplementary Table S11). Lower educational attainment was also associated with strong positive responses, for 4 out of 6 healthcare preference items. Older age, female sex and lower educational attainment were each associated with higher odds of selecting strong negative scores for some items (Supplementary Table S12).

## Discussion

Six new measures of general healthcare preferences were administered to people aged 50 and over in England: risk aversion, future orientation, quality or length of life, body function compared to looks, openness to experimental treatments, and preference to leave treatment decisions to a doctor. Reported healthcare preferences varied between individuals as expected, but there were some systematic associations with age and sex which may be a useful aid to the elicitation of healthcare preferences in a busy clinical environment. Notably, women were less likely than men to want to leave treatment decisions to doctors, and more likely to prioritise quality of life over length of life. Participants aged over 75 years were more likely to want to leave treatment decisions to doctors, and more likely to want to avoid risks. The relatively low correlations we observe between different dimensions of healthcare preferences suggest that these domains represent distinct aspects of patient priorities, each with different effects on patient engagement strategies.

This study is the first to explore general healthcare preferences in older adults, and the findings align with studies of preferences in specific health conditions. A recent systematic review of health preferences in older adults with cancer found that older adults prioritised current quality of life over length of life, and relied heavily on expert advice.[19] A recent USA study found that older age and female sex were associated with prioritising quality of life over length.[20] A 2010 study from the USA found that women with chronic disease (hypertension, diabetes, myocardial infarction, congestive heart failure, and depression) were 44% more likely to desire active participation in medical decision-making compared to men.[21]

A major strength of the study is the large, representative ELSA sample, which supports generalisability of findings to older adults in England. The healthcare preference items are unique to ELSA and have not yet been asked in other surveys. Limitations include that the questions ask about hypothetical preferences, and these preferences have not been tested with real health care decisions. Other unmeasured aspects of preferences may also be important for healthcare decisions.

### Implications for research and practice

Further research is needed to test these general healthcare preferences in other populations, and with real health care decisions. Healthcare preferences are not static but can evolve over time as patients’ conditions and life circumstances change.(2) Further longitudinal research should explore how healthcare preferences evolve over time, particularly with respect to changes in health status or life circumstances, and how often they should be reassessed to ensure that medical decisions remain aligned with patient preferences. Including more domains of healthcare preferences, such as familial considerations, could provide a more holistic understanding of patient-centred care and support tailored approaches that respect patient preferences.

These findings have important implications for clinical practice. Whilst healthcare preferences vary between individuals and cannot be assumed to hold across demographic groups, there are some general preference traits by age and sex and education that may be useful to clinicians seeking to elicit patients’ preferences within the constraints of a short consultation. Shared decision making tools such as decision aids can facilitate patient-provider conversations that reveal individual preferences, particularly where patients may have distinct perspectives on treatment risks, benefits, and autonomy.

## Conclusions

The new information presented here about associations between healthcare preferences and patient characteristics offers potential for the quicker elicitation of individual preferences in medical practice. These associations cannot replace assessment of preferences with individual patients, and further research and testing is needed, but cautious use of these results to facilitate the elicitation of health preferences is likely to lead to better health outcomes and greater patient satisfaction with care.

## Data Availability

Data are available on UKDS: SN 5050 English Longitudinal Study of Ageing: Waves 0-11, 1998-2024. https://datacatalogue.ukdataservice.ac.uk/studies/study/5050#details10.5255/UKDA-SN-5050-35 Citation: Banks, J., Batty, G. David, Breedvelt, J., Coughlin, K., Crawford, R., Marmot, M., Nazroo, J., Oldfield, Z., Steel, N., Steptoe, A., Wood, M., Zaninotto, P. (2025). English Longitudinal Study of Ageing: Waves 0-11, 1998-2024. [data collection]. 48th Edition. UK Data Service. SN: 5050, DOI: http://doi.org/10.5255/UKDA-SN-5050-35

https://datacatalogue.ukdataservice.ac.uk/studies/study/5050#doi

## Supporting information captions (Supplementary Tables 401 S1-S12).

**Table S1. Associations between covariates and the health preference item ‘avoid risks’, English Longitudinal Study of Ageing wave 8 (linear regression coefficients)**

**Table S2. Associations between covariates and the health preference item ‘live for the future’ (linear regression coefficients)**

**Table S3. Associations between covariates and the health preference item ‘length of life’ (linear regression coefficients)**

**Table S4. Associations between covariates and the health preference item ‘body function’ (linear regression coefficients)**

**Table S5. Associations between covariates and the health preference item ‘avoid experimental treatments’ (linear regression coefficients)**

**Table S6. Associations between covariates and the health preference item ‘leave treatment decisions to doctor’ (linear regression coefficients)**

**Table S7. Percentage of variance* in health preference items explained by covariate clusters: comparison of final sample and restricted sample, ELSA Wave 8. All analyses use weighted samples (linear regression).**

**Table S8. Associations between participant characteristics and health preferences (linear regression coefficients). Restricted sample with no missing data.**

**Table S9. Associations between participant characteristics and health preferences with all responses with a score of 5 excluded (linear regression coefficients)**

**Table S10. The likelihood of responding with score “5” vs “not 5” (logistic regression odds ratios)**

**Table S11. The likelihood of responding with strong positive scores “≥6” vs “≤5” (logistic regression odds ratios)**

**Table S12. The likelihood of responding with strong negative scores “<5” vs “≥5” (logistic regression odds ratios)**

